# Manual sub-segmentation of the cererbellum

**DOI:** 10.1101/2022.05.09.22274814

**Authors:** Jennifer Faber, Lea-Sophie Heinz, Dagmar Timmann, Thomas M. Ernst, Katerina Deike-Hofmann, Thomas Klockgether

## Abstract

1

**Abstract:** The protocol was developed in the context of generating manual sub-segmentations of the cerebellum to use as ground truth for training, validation and testing of a neural network. In this manuscript we provide methodological considerations and results of our manual sub-segmentation as well as a detailed description for manual sub-segmentation of the cerebellum into 10 hemispheric lobules and 5 vermal sub-segments as well as two cerebellar white matter labels based on a T1 weighted (T1w) MRI to foster reproducibility.

## 1.2 Anatomical nomenclature

**Figure 1:**
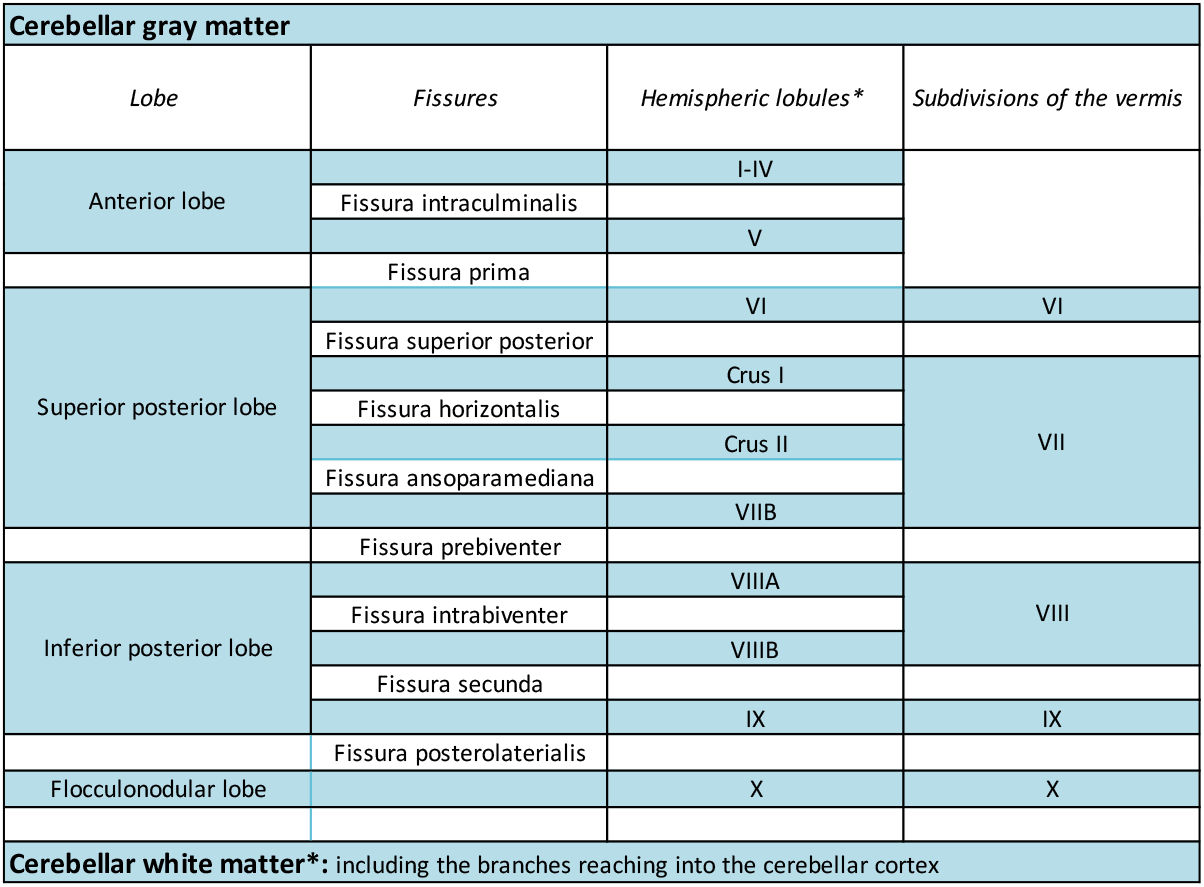
Nomenclature of the cerebellar sub-structures segmented in this protocol. *The hemispheric lobules as well as the cerebellar white matter are segmented separately for the left and right hemipshere. The nomenclature of the cerebllar gray matter is according to Schmahmann et al. [1]. Since all labels are disjoint, they can be combined by simple addition to form the cerebellar lobes, the vermis, or the entire gray matter of the cerebellar cortex.

## 1.3 Introduction

The cerebellum is located in the posterior cranial fossa. It has a remarkably higher cell density than the cerebrum and, although it is only 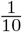 the size of the normal adult brain, contains 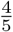 of all neurons and, for its size, multiple times the surface area of the cerebrum.[2, 3] Computed tomography (CT) lags far behind magnetic resonance imaging (MRI) in its ability to capture detail of the cerebellar macroscopic structure. Anatomically, the cerebellum is divided into the anterior lobe, the superior and inferior posterior lobe and the flocculonodular lobe. In its posterior parts the midline vermis can be distinguished from the lateral hemispheres. The next smaller anatomical subdivision of the cerebellar lobes are ten lobules I-X, of which two, namely VII and VIII, are again further sub-divided into two parts in themselves. In general, a lobule can consist of one or more folia, i.e., the gray matter cortex surrounding a branch of white matter emanating from the medullary corpus of the cerebellum. The midline worm can be subdivided accordingly, but there are only very limited macroscopic landmarks for each subdivision, so that the subdivision largely corresponds to the extrapolation of the hemispheric lobule boundaries. In general, the cerebellum shows a high morphologic variability of its anatomical subdivisions.

In this protocol we provide a detailed description for manual sub-segmentation of the cerebellum into 10 hemispheric lobules and 5 vermal sub-segments as well as two cerebellar white matter labels based on a T1 weighted (T1w) MRI as well as methodological considerations and results of our manual sub-segmentation process. We established the protocol for an in-house manual cerebellar sub-segmentation of 30 MRI scans that should serve as ground truth for an automated segmentation. All of these scans were T1-weighted MPRAGE with an 1mm isotropic resolution acquired with a fieldstrength of 3 Tesla on SIEMENS scanners. We continuously updated the written record of the protocol during the segmentation process as well as based on feedback from independent colleagues who used the protocol to replicate the segmentations, in order to optimize the description of the most difficult subdivisions and make it as clear as possible.

### Cerebellar Cortex

As mentioned above, the cerebellar cortex is hierarchically divided into lobes, followed by further subdivisions into lobules. While anatomic landmarks are clearly identifiable and distinct for most sub-segments, some cerebellar sub-structures cannot be adequately and consistently imaged with conventional MRI because of their small size. Therefore, it was necessary to make some determinations in advance, which basically refer to the lobules of the anterior lobe and the sub-segmentation of the vermis. The anterior lobe was subdivided into lobules I-IV as an aggregated volume and lobule V. We did not further subdivide lobules I-IV. Anatomically, the precentral fissure separates the hemipheric lobules I-II from lobule III and the preculminate fissure separates lobule III from lobule IV. Lobule III has a semilunar form and in most cases the first distinct folium not attached to the superior medullary velum. However it is often at least partially obscured by lobule IV. With the spatially unbiased atlas template of the cerebellum and brainstem, the group of Diedrichsen et al.[4] provided a probabilistic atlas of cerebellar anatomy to assign locations to different cerebellar lobules and deep cerebellar nuclei. Here, lobules I-IV are aggregated, too, according to their strong functional relation, being primarely involved in motor function of the upper limb as shown in various functional MRI studies [5–9]. Due to the partially vague fissures together with the close functional context, we also decided not to subdivide lobules I-IV for now, although there are examples of further subdivisions of the anterior lobe from other groups [10, 11]. Regarding the midline vermis, vermal branches can be traced and separated in the anterior lobe only on a microscopic level. However, this is not possible on a macroscopirc level.[1, 12] Consequently and in accordance with other procotols, the vermis was segmented in the posterior part of the cerebellum, corresponding to the lobules VI to X.[1, 4, 10, 11] In theory, the vermis is separated from the cerebellar hemispheres by the paravermian sulcus. However, this may be obscured by the indentation resulting from the course of the superior cerebellar artery. In addition, oblique fissures, in particular the preculminate and intraculminate fissure, make a definite assignment difficult. We decided to subdevide the vermis in sub-segments according the hemispheric lobules VI, VII, VIII, IX and X as a pragmatic compromise between a segmentation only in its entirety and a sub-segmentation according to all hemisperic lobules, that would include subdivision of lobule VII into Crus I, Crus II and VIIB as well as the subdivision of lobule VIII into VIIIA and VIIIB, resulting in very small volumes of partially only a few voxels.

### Cerebellar white matter

The consistent segmentation of the cerbellar white matter is important in demarcation from both the cerebellar cortex and the brainstem. Regarding the boundary to the cerebellar gray matter, in contrast to the cerebrum, the white matter of the cerebellum reaches into the cerebellar cortex with widely branched ramifications. Regarding the boundary to the brainstem it has to be noted, that anatomically, the paired superior, middle and inferior cerebellar peduncles are distinguished containing the efferent and afferent fibre bundles connecting the cerebellum with the midbrain, pons and medulla oblongata respectively. The paired superior cerebellar peduncles can be easily identified as separate structures, while a separation of the middle and inferior cerebellar peduncles is not possible on a T1w MRI. Consequently, the specifications for the boundary of cerebellar white matter need to be clear and reproducible, regarding the branching into the cortex folia as well as towards the brainstem. The delineation of white matter branches down to the finest branches that reach into the gray matter cortex is limited by the resolution of the MRI scan. We labeled voxels of white matter reaching into the cerebellar cortex as white matter as long as one voxel shares a common face with a directly adjacent voxel that has already been assigned to white matter (which corresponds to a common edge in a 2D view). For the boundary of cerebellar white matter towards the brainstem we selected the superior and inferior point where cortex gray matter touches white matter in sagittal slices as anatomical landmarks and draw a connection line between them. Finally, all of these connection lines were corrected in the axial and coronal views in order to achieve a smoothed surface.

We recommend to study the Three-Dimensional MRI Atlas of the Human Cerebellum in Proportional Stereotaxic Space"[1] for anyone intending a manual sub-segmentation of the cerebellum. Here, anatomical slices of the cerebellum are directly compared with corresponding slices of MR images, thus facilitating the identification of anatomical landmarks. In addition, we recommend to also have a look Bogovic et al.[10] and Park et al.[11]. In our protocol we are following the nomenclature introduced by Schmahmann et al. [1] An overview of the segmented structures is given in Section 1.2.

In the section on methods (Section 1.4) we describe basic instructions. By reporting the results of comparisons between trained raters we provide indications of which structures were particularly prone to uncertainty in the identification of anatomical boundaries (Section 1.5). In the section on the segmentation protocol (Section 2), all hierarchical steps for a full segmentation of the cerebellum are listed. We have taken particular effort to describe each segmentation step as accurately and unambiguously as possible, with notations of particular pitfalls and difficulties, as well as common anatomical variations. We welcome feedback on where the protocol can be further improved.

## 1.4 Methods

### 1.4.1 General recommendations for the manual segmentation

#### Preconditions and technical equipment

Prior to segmentation, all scans need to be checked for image quality in order to exclude images that are unsuitable for manual segmentation due to artifacts, e.g. motion artifacts, or an insufficient coverage of the entire cerebellum. Image contrast of the T1w MRI should be optimized first and needs to be kept constant for all segmentation steps. For our protocol we used ITK-SNAP, version 3.6.0, to manually delineate the cerebellar sub-segments (http://www.itksnap.org) [13]. ITK-SNAP is an open-source application, that allows to mark different structures with a specific color label for each voxel of an MRI image and runs in a Windows as well as Linux or iOS environment. ITK-SNAP offers to display labels as a three-dimensional reconstruction in addition to the axial, coronal and sagittal view, which can be helpful to doublecheck the integrity and consistency of the outer surface of a drawn 3D-structure. There are other freely available software solutions that can also be used, e.g. from *FreeSurfer* (http://surfer.nmr.mgh.harvard.edu) or FSL (https://fsl.fmrib.ox.ac.uk/fsl/fslwiki/FSLeyes). We recommend to double check the correct assignment of left and right to avoid unintended flipping. We used a pen display for drawing. In our color coding scheme we tried to avoid similar colors of directly adjacent structures Figure 2.

**Figure 2:**
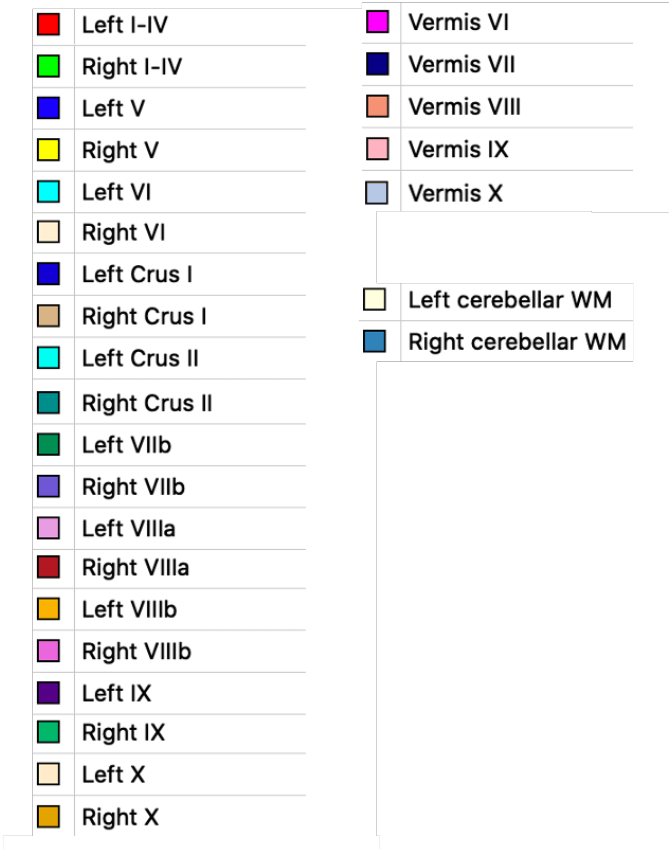
Look up table (LUT) of the color scheme. Please note: Figures in this Protocol might differ from the LUT. The colors were chosen in such a way that the separation of the represented structures was the best possible for each Figure separately.

#### Use of pre-segmentations

Even though manual segmentation is very time consuming we do not recommend to use any pre-segmentations in general. The advantage of using automated pre-segmentations is a faster segmentation process, since in particular planar coloring of larger structures is time consuming and might be reduced at least for the inner areas of sub-segments. But, this approach runs the risk of subconsciously following the pre-determined boundaries set by the automated method. If you are a newbie starting with cerebellar sub-segmentation, we strongly recommend to use native MRI. Trained users may be able to use such pre-labeled sub-segmentations, as long as they review them very critically. Again, we recommend defining the lobe boundaries on a native MRI before looking at the pre-labels, and only using the pre-labels when painting out the inner regions of the sub-structures. Since the ground truth of *CerebNet* has been labeled according to the protocol presented here, it makes sense to use *CerebNet* for the automated pre-segmentation of sub-structures. We would like to point out that such a procedure does not follow the hierarchical steps of sub-segmentation. This protocol contains 6 hierarchical steps from large to more and more detailed sub-structures of the cerebellum. The first step is the drawing of the entire cerebellum. As a compromise to reduce the amount of time for manual segmentation, the use of a pre-determined cerebellar mask for this first step seems acceptable to us also for untrained raters. This automated cerebellar mask needs to be critically reviewed and corrected. However, we consider the risk of mislead manual segmentations due to the pre-segmentation to be relatively low, while the potential of saving time is relatively large. Beside tools for cerebellar sub-segmentation such as *CerebNet, ACAPULCO* or *CERES*, also e.g. *FreeSurfer, FastSurfer* as well as the *SUIT* toolbox provide cerebellar masks without sub-segmentation, that can be used as a starting point. Please make sure, to carefully correct not only ‘over-segmentations’ that include non-cerebllar structures, such as cranial nerves, venes or dura, but also to correct ‘under-segmentations’ not labeling the periphery of the cerebellum, e.g. at the apex of its triangle-like parts in the midline.

#### Anatomical reference

The atlas “Three-Dimensional MRI Atlas of the Human Cerebellum in Proportional Stereotaxic Space” by Jeremy Schmahmann is a key reference, which we highly recommend [1]. The nomenclature introduced by Schmahmann has become common. In the atlas, MRI images are presented alongside anatomical slices. This is very helpful and provides orientation for cerebellar anatomical sub-structures exceeding the location of the deep cerebellar nuclei that cannot be captured on T1w MRI. However, it does not cover different subjects and therefore has a shortcoming in terms of representing intraindividual variability. It is important keeping this in mind, if the actual scan one is working on, does not look exactly the same as in the atlas. In such aberrant cases, it is not possible to simply copy-and-paste from the atlas. In these cases, the course of the fissures must be traced as precisely as possible in the present MRI in order to correctly define and delineate the cerebellar structures.

#### Use of the different views - axial, sagittal, coronal - and a 3D model

Segmentations of boundaries in the cerebellum are always performed in the view in which the respective structure can best be followed and thus depends on its location and spatial course. For the overall segmentation of the entire cerebellum the axial view is a got starting point, while most fissures that separate adjacent lobules can best be followed in the sagittal view. However, often two or even all three views are needed to complete even the first “draft” of the outer boundary of a structure. After each drawing in one view, the sub-segments must be checked and corrected in the other views. It is important to repeat this iteratively and conclude with a final inspection of each cerebellar structure in all views: sagittal, coronal, and axial. It is important to keep in mind, that the underlying “real” anatomy has smooth boundaries. Therefore, we also recommend to check the surface in a 3D model (see Figure 3) for any obvious inconsistencies.

**Figure 3:**
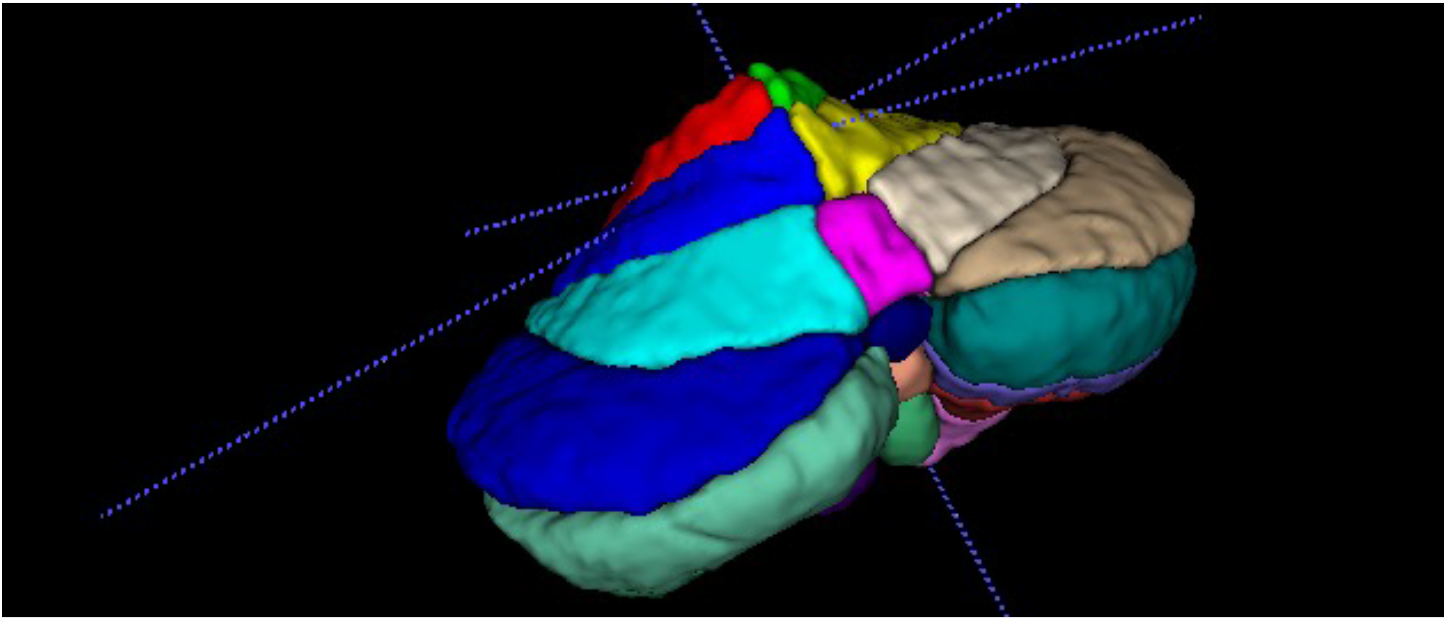
Example of a completely sub-segmented cerebellum in the 3D model in dorsal view. You can see the superficial contours of all labels. Each label should also be displayed separately to doublecheck the outer boundaries.

#### Hierarchical approach

In order to avoid the accidental labeling of voxels e.g. without reference to the cerebellum, we recommend to follow our hierarchical approach starting from large to increasingly detailed cerebellar sub-structures (e.g. entire cerebellum >cerbellar cortex >lobes >lobules >detailed WM) and recommend always to ‘paint over’ using the next larger structure as a ‘background label’. By doing so, misclassifications, e.g. drawing of empty voxels by the current ‘foreground label’ or ‘active label’, is technically not possible. In other words, if you have in the first step segmented the entire cerebellum, use this as the background label for the lobes. Specifically, this means that only voxels which have already been classified as ‘cerebellum’ are now assigned to respective lobes. Thus, whereas in the first step of segmenting the entire cerebellum one had to focus very carefully on the outer boundary (separating the cerebellum from cranial nerves, sinus etc.), one can now concentrate on the current challenge of a precise delineation of the fissures representing the intra-cortical boundaries that separate the different lobes and one can simply paint over the outer surface since it is ensured, that only the ‘background voxels’ already assigned to the cerebellum are coloured. Finally, it has to be assured that each voxel is uniquely assigned to a cerebellar sub-structure of the *final* 27 sub-structures, thus resulting in a disjoint segmentation of the cerebellum. In other words, there should be no voxel labeled as “inferior posterior lobe” not assigned to either VIIIA, VIIIB, or IX. Usually, respective software solutions use a numeric index for each label. As a practical advice, we strongly recommend to use different indices for the large structures (such as cerebellar gray and white matter, cerebellar lobes and overall vermis, according to STEP 1 - 4) than those numeric indeces that will later be used for any of the final 27 sub-structures. This also simplifies a final check whether all voxels have been assigned to one of the 27 final sub-structures.

#### Documentation

For each segmentation a documentation file should be set up, e.g. an excel sheet, in which technical data as well as notes from the labeling process are recorded. Technical data should include contrast adjustment data, image intensities, image and voxel size as well as scanner information, including vendor and field strength, and sequence parameters. Furhtermore, all steps and structures should be listed and either marked as “done” or commented on in detail in case of uncertainties or discrepancies. It is advisable to have a second look at all segmentation steps that were found to be problematic after a while.

#### Rater and consensus ratin

Raters should be trained on working with 3D MRI. We strongly recommend to have a team of raters to discuss ambiguous and border cases. We recommend untrained persons to re-do their first scans after having gained some more practical experience. We proceeded as follows: The challenging segmentation of the cerebellar grey matter into the lobules was performed independently for all scans by two raters. Subsequently, those two segmentations were manually compared for each subject and each single discrepancy, e.g. the assignment of a single branch to Crus II by rater 1 versus Crus I by rater 2, was reviewed and discussed in detail. With the inclusion of the expertise of a neuroradiologist, consensus voting could thereby be achieved for each individual structure.

## 1.5 Results of the comparison between raters

We have labeled 30 scans. All participants gave their written informed consent for the use of the MRI data.The 30 subjects included healthy controls as well as pre-ataxic spinocerebellar ataxia type 3 mutation carriers with only subtle cerebellar atrophy as well as symptomatic patients with various degrees of mild to severe cerebellar atrophy. The cerebellar gray matter was labeled by 2 raters independently. In total, we observerd deviating anatomical boundaries in 67 volumes. This was related the following structures in descending frequency: Fissura ansoparamediana (*N* = 27), Fissura prebiventer (*N* = 11), Fissura intraculminalis (*N* = 7), Fissura intrabiventer (*N* = 6), boundary of Lobulx X (*N* = 5), Fissura secunda (*N* = 4), Fissura prima (*N* = 3), Fissura horizontalis (*N* = 1), Fissura posterolateralis (*N* = 1), subsegmentation of vermis VI (*N* = 1) and vermis VIIIB (*N* = 1). Numbers of deviating anatomical boundardies were comparable between healthy controls and pre-ataxic spinocerebellar ataxia type 3 mutation carriers that show only very subtle cerebellar atrophy (controls: 18 out of 250 volumes; pre-ataxic mutation carriers: 17 out of 275 volumes) while in symptomatic patients numbers of deviating anatomical boundaries were much higher (32 out of 225 volumes). Thus, atrophy obviously leads to a higher rate of uncertainty in identifying the correct fissures and thereby in defining the anatomical boundaries.

## 2 Segmentation of the cerebellum

### General remarks

This protocol uses a hierarchical approach from large to more and more detailed sub-structures of the cerebellum. The final goal is a disjoint subdivision of the cerebellum into the following 27 cerebellar sub-structure lables: *left and right I-IV, left and right V, left and right VI, left and right Crus I, left and right Crus II, left and right VIIB, left and right VIIIA, left and right VIIIB, left and right IX, left and right X, vermis VI, vermis VII, vermis VIII, vermis IX* and *vermis X* as well as *left and right cerebellar white matter*. Label indices for the large structures (such as cerebellar gray and white matter, cerebellar lobes and overall vermis, according to STEP 1 - 4) should be different from those of the final 27 sub-structures. For each substructure, we provide recommendations on which view is best to start segmenting. After each drawing in one view, the sub-segments must be checked and corrected in the other views. It is important to repeat this iteratively and conclude with a final inspection of each cerebellar structure in all views: sagittal, coronal, and axial as well as the 3D model, see also Section 1.4.1. We consider this final check for each sub-structure a basic process and do not repeat it at the end of each of the following segmentation steps.

#### 2.1 STEP 1: Overall segmentation of the cerebellum

We recommend to start with the outer cerebellar boundary of the cerebellar gray matter. The cerebellar cortex is separated from the cerebrospinal fluid and the adjacent tissues such as the cerebrum, cranial nerves, venes/sinus, brainstem and meninges. The dura mater is inferior of the tentorium cerebelli and does not belong to the cerebellar gray matter. As mentioned above, we do not recommend to use any pre-segmentation, but consider the use of a segmentation for the entire cerebellum as a starting point for the first step to be acceptable (Section 1.4.1). Pre-segmentations should be critically reviewed and over- as well as under-segmentations need to be corrected. Especially the outer boundary of the superior posterior cerebellum is often over-segmented by automated methods, including sinus, tentorium and adjacent cerebral parts (Figure 4 shows an example) as well as cranial nerves, in particular close to lobule X (Figure 20). As for all segmentations we recommend to iteratively correct each drawn boundary in all views. To segment the outer boundary of the cerebellar cortex we recommend to start in the axial view going from top to bottom. Second, use the sagittal view for a correction, in particular of the superior and dorsal expansion and subsequently, the correction of the inferior and anterior outer cerebellar cortex boundary. Finally, verify the outer cerebellar cortex boundary in the coronal view and again in the axial and sagittal view as well as in the 3D model to check the overall outer surface smoothness. The following structures need to be carefully doublechecked as non-cerebellar structures: the tentorium, the most inferior parts of the cerebrum, cerebral sinus and cranial nerves. In this step the entire cerebellar white matter (WM) is segmented, too (see Figure 4). The exact boundary between cerebellum and brainstem is skipped in this step. We recommend to segment the white matter in the middle cerebellar peduncle to some extend towards the brainstem. By doing so, it will be more easy to just crop protruding parts when in comes to the exact and consistent segmentation of the WM boundary in STEP 6 Section 2.6.

**Figure 4:**
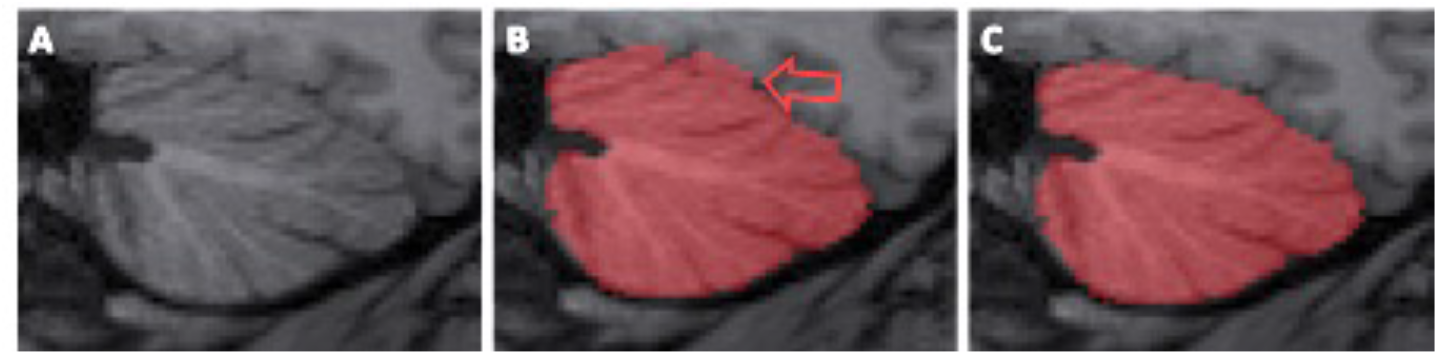
Example for the oversegmentation of the entire cerebellum including surrounding tissue and cerebrospinal fluid. The native MRI of a sagittal slice of the left hemisphere is shown in (A). Here, the dorsal, overshooting presegmentation exceeded the cerebellar anatomic boundary (B, marked with the red arrow) and was manually corrected (C).

#### 2.2 STEP 2: Segmentation of the Lobes

In this step the next level of cerebellar sub-structures is segmented: The anterior lobe, the superior-posterior lobe and inferior-posterior lobe. For the segmentation of the flocculonodular lobe, lobule X, we refer to the corresponding paragraph in STEP 4 Section 2.4.10. The fissures relevant to this step are the primary fissure, which separates the lobules V and VI, and the prebiventer fissure or Fissura prepyramidalis, which separates the lobules VIIB and VIIIA. In the midsagittal plane those two fissures are well recognizable. Their spatial course in lateral direction through the hemispheres can be followed and traced slice by slice in the sagittal view. Finally, the segmentation of the lobes has to be cross-checked and if necessary corrected in the coronal and axial views, see Figure 5. In this step a preliminary boundary of cerebellar cortex towards the cerebellar white matter is drawn. Any branching ramifications of white matter into the cerebellar cortex are ignored and only a straight boundary at the basis of each branch is drawn in form of a straight connection line in form between the opposing limits of gray matter (Figure 5). The detailed segmentation of the white matter is done in STEP 6 (Section 2.6).

**Figure 5:**
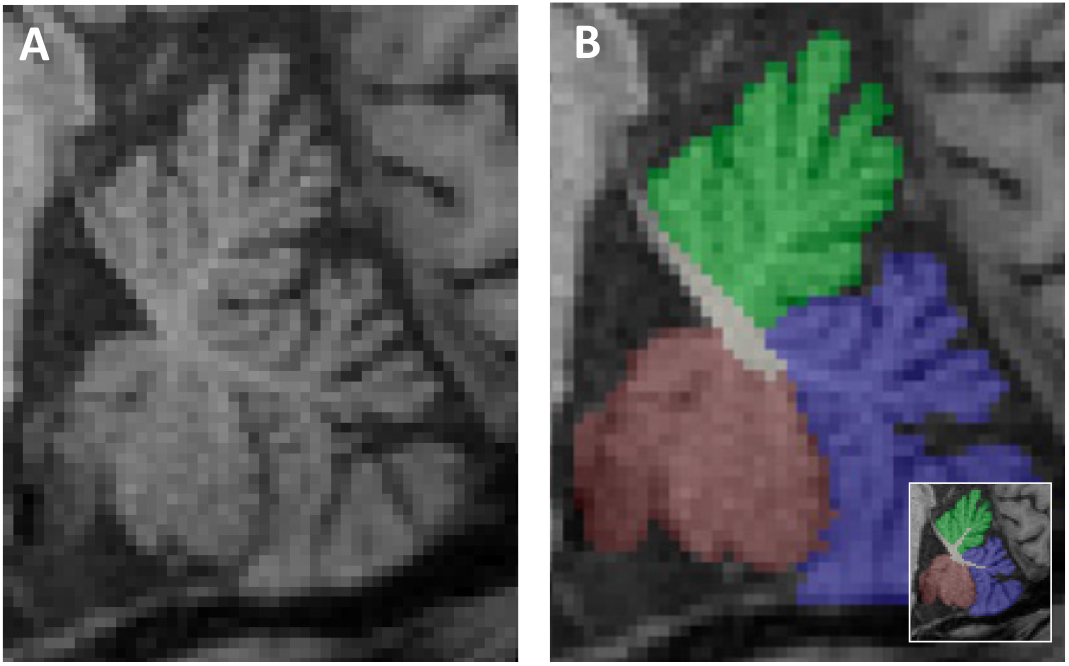
Segmentation of the lobes. Anterior lobe (green), posterior superior lobe (blue) and posterior inferior lobe (wine red) projected (B) onto a sagittal slice close to the midsagittal plane (A). Please note: White matter branches projecting into the cerebellar cortex are ignored in this step. For comparison, the final segmentation, including the fine-grained boundary of the white matter that captures the branched ramifications into the cortex, is shown in the lower right corner of (B).

#### 2.3 STEP 3: Segmentation of the overall Vermis

The vermis (Latin for worm) forms the narrow midline part between the two cerebellar hemispheres bordering on the medullary corpus and the fourth ventricle. The vermis cannot be macroscopically defined in the anterior lobe [1, 12]. Consequently, the vermis is only segmented in the posterior parts of the cerebellum. First, the vermis is segmented in its entirety. Finally, the vermis is subdivided in analogy to the hemispheric lobules in the posterior cerebellum. In this STEP 3 we will focus on the segmentation of the overall vermis in its entirety. The symmetrical outline of the vermis is best seen in the coronal plane, as shown in Figure 6. Best distinguishable is the boundary of the vermis to the surrounding tissue in its most ventral and most dorsal parts. In the dorsal parts, the vermis appears as a rounded, longish structure between the hemispheres, most of which is inferior and posterior to the medullary corpus. In the ventral sections the vermis impresses as a triangular structure with the apex of the triangle pointing inferiorly. The dorsal extent of the vermis can best be followed in the axial plane, starting with the delineation of the vermis adjacent to the hemispheric lobules VI. Here, the vermis presents roundly shaped in the midline extending (in anterior-posterior direction) over the whole length of the lobule VI. The ventral portion of the vermis can be found in the axial plane in the more inferior sections between the cerebellar tonsils. The portion of vermis belonging to lobule X seems to project into the fourth ventricle, while vermis IX impresses like an arrowhead. The outlines of the vermis corresponding to the more inferior part of VII (corresponding to hemispheric VIIB) and VIII imposes arched, where the arc opens ventrally, while the more superior part of the vermis VII (corresponding to the hemispheric Crus II) appears more like a triangle. The ventral and dorsal parts of the entire vermis are finally connected in their middle. See Figure 6. The vermal sub-segmentation corresponding to the hemispheric lobules will be completed in a later step (Section 2.5).

**Figure 6:**
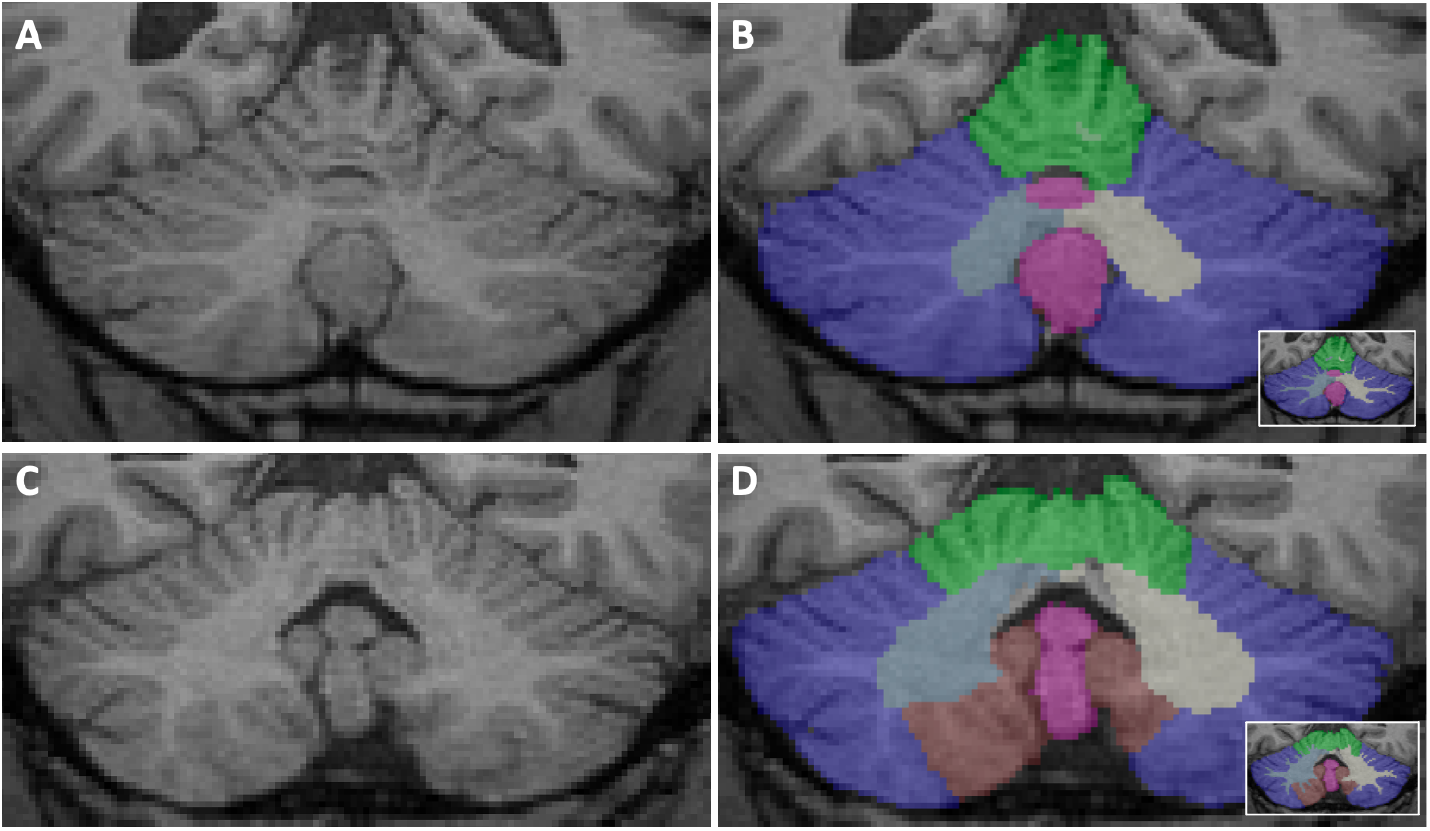
Example of segmentations of the vermis (pink) as well as the cerebellar lobes on two coronal slices. (A) and (B) showing a coronal plane where the vermis forms an inferiorly rounded elongated structure in the dorsal part (colored in pink). In (C) and (D) the vermis can be seen in the more ventral section with a triangular and longish shape (colored in pink). The anterior lobe (green), posterior superior lobe (blue) and posterior inferior lobe (wine red) are shown for orientation. For comparison, the final segmentation with white matter boundary capturing the branching into the cerebellar cortex is shown in the right bottom corner of (B) and (D).

#### 2.4 STEP 4: Segmentation of the lobules

##### 2.4.1 Lobules I-IV

The lobules I-IV are the most superior group of lobules which form together with lobule V the anterior lobe. Lobules I and II, which are often not differentiable as single structures, are close to the superior cerebellar peduncles. They show a large variability between individuals, which has a strong impact on volume differences due to the small size of the structure (see also [11]). Dorsal to lobule II, separated by the preculminate fissure, follows lobule III, which arises from a narrow branch of white matter. Lobule III has a semilunar form and in most cases the first distinct folium not attached to the superior medullary velum. Anatomically, the preculminate fissure separates it from lobule IV, however, lobule III is often at least partially obscured by lobule IV. Lobules I-IV are segmented together as an aggregated structure. Consequently, the relevant fissure that needs to be identified is the intraculminal fissure, separating lobule IV from lobule V. Lobules IV and V form a tree-like structure and share the same branch of white matter, which can best be seen in the midsagittal plane. Notably, the common portion of the branch is already divided, before a separation through the intraculminal fissure into the two lobules can be clearly seen on the MRI. The separating intraculminal fissure between lobules IV and V is best determined in the slice of the midsagittal plane and a few slices laterally. From there on it needs to be followed laterally to the sides. It should be traced switching between the sagittal and coronal view. See Figure 7.

**Figure 7:**
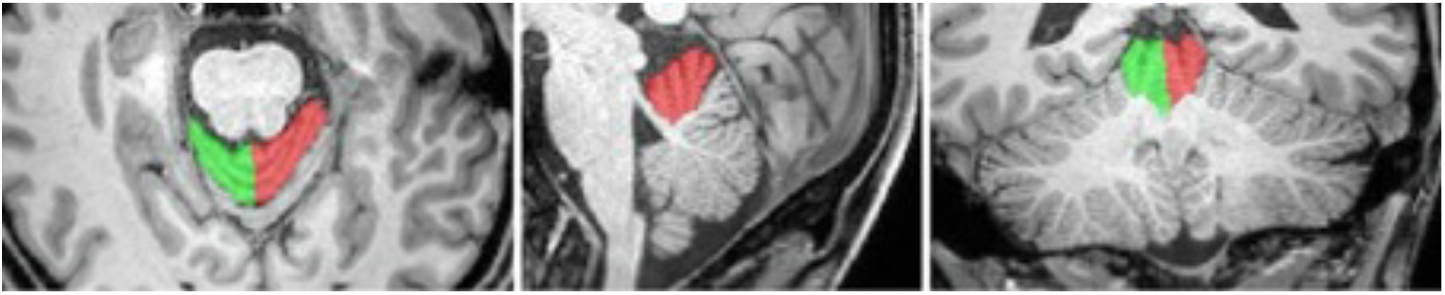
Lobule I-IV in axial, sagittal and coronal view. Red: left I-IV. Green: right I-IV.

##### 2.4.2 Lobule V

lobule V usually has 2 to 3 folia, and thus often one more folium than lobule IV. It is separated from lobule VI by the primary fissure, which appears most prominently in the sagittal midline and has already been identified in STEP 2 Section 2.2, Segmentation of lobes. Figure 8 showes Lobule V in both cerebellar hemispheres.

**Figure 8:**
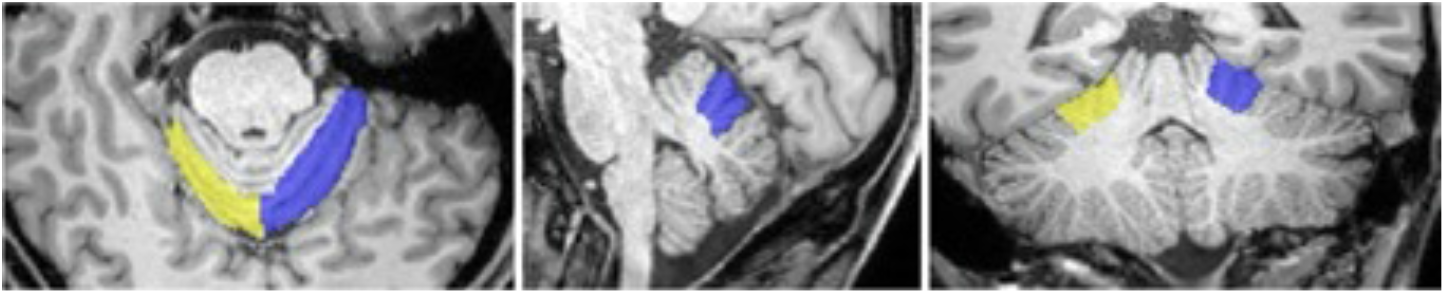
Lobule V in axial, sagittal and coronal view. Blue: left V. Yellow: right V.

##### 2.4.3 Lobule VI

Lobule VI has 2 to 3 folia and is bounded posteriorly and inferiorly by the superior posterior fissure. In the midline and a few millimeters further lateral, lobule VI often borders directly adjacent to Crus II, as Crus I regularly narrows medially and may eventually ‘disappear’. To identify the fissure between lobule VI and Crus I, the sagittal plane about 15 mm lateral to the midline is used. From here, the fissure is tracked laterally throughout the hemispheres. Afterwards the missing medial portion is completed. See Figure 9. The superior posterior fissure can be traced and controlled in the axial plane throughout the entire hemisphere (see left image in Figure 9).

**Figure 9:**
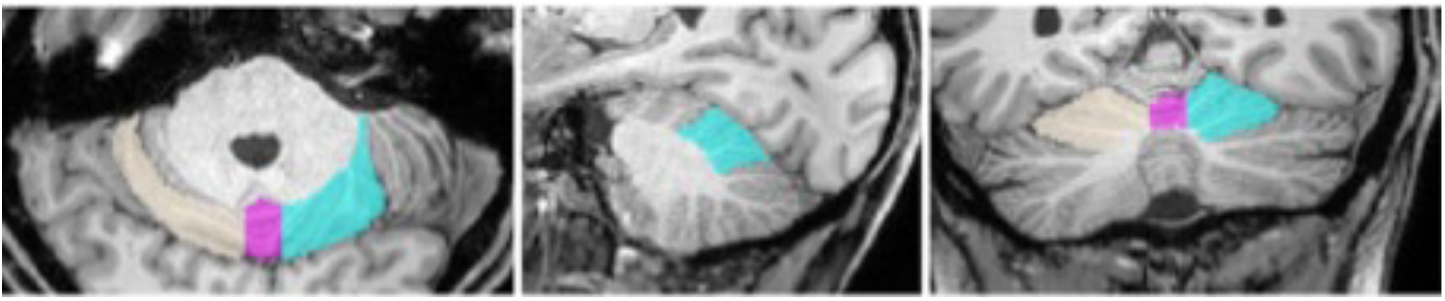
Lobule VI in axial, sagittal and coronal view. Turquoise: left VI. Beige: right VI. Pink: Vermis.

##### 2.4.4 Crus I

Crus I is a large lobule and arises from only one branch, which usually appears laterally of the midsagittal plane. In the periphery, the lobule further enlarges and reaches the largest extent of all lobules in the lateral periphery. It is separated from Crus II by the horizontal fissure, which can be detected very well in the dorsal slices using the coronal view. Starting from here, the fissure representing the lobules boundary can be traced. Differences in intensity are also often clearly visible at the axial view, thus the axial view is continuously used for doublechecking and re-drawing if necessary. See Figure 10

**Figure 10:**
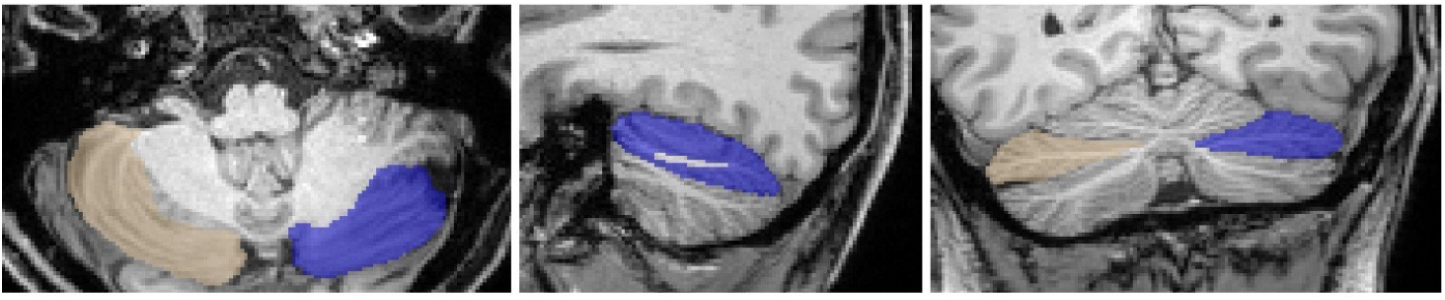
Crus I in axial, sagittal and coronal view. Blue: left Crus I. Light brown: right Crus I.

##### 2.4.5 Crus II

Crus II is located inferior to the horizontal fissure and also originates from one branch of white matter, but may vary in the number of folia and size. Crus II often shows an asymmetry, being more prominent on the right side than in the left hemisphere. Thus, the two limiting fissures (Fissure horizontalis and Fissura ansoparamediana) often merge laterally in the left, but not in the right hemisphere. Consequently, this results in the triangular shape of Crus II on the left side (see also [1]). In addition, on the left side, Crus II often shares the same branch of white matter with lobule VIIB (see also [11]). Crus II is separated from lobule VIIB by the fissura ansoparamediana, which is best seen in sagittal view about halfway between the midline and the lateral end of the cerebellum and should be followed from there in a medial and lateral direction (see Figure 11, middle picture showing the sagittal view). The coronal view is well suited to track this fissure. In the axial plane, where lobule VIIB often surrounds from anterior and ventral the lateral portion of Crus II, tracing the fissure becomes more difficult as the boundaries often appear somehow blurred.

**Figure 11:**
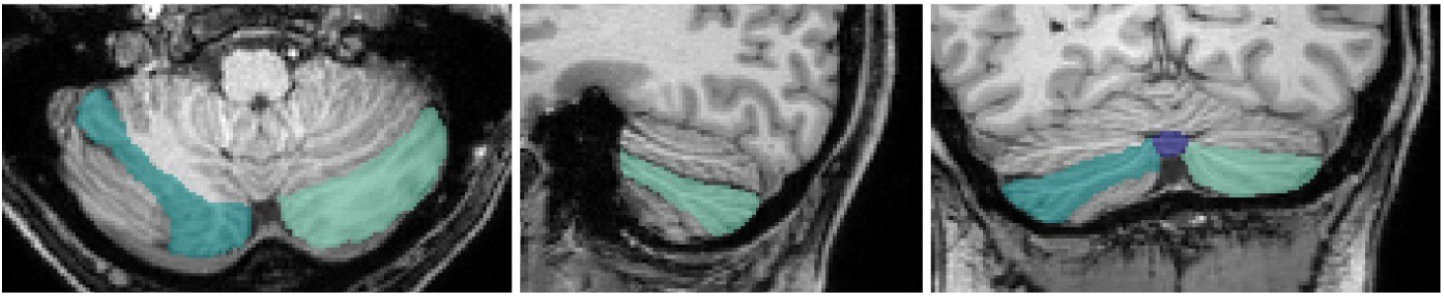
Crus II in axial, sagittal and coronal view. Light green: left Crus II. Dark green: right Crus II.

##### 2.4.6 Lobule VIIB

Lobule VIIB is separated from lobule VIIIA by the fissura prebiventer or prepyramidalis that has already been defined, see Figure 12.

**Figure 12:**
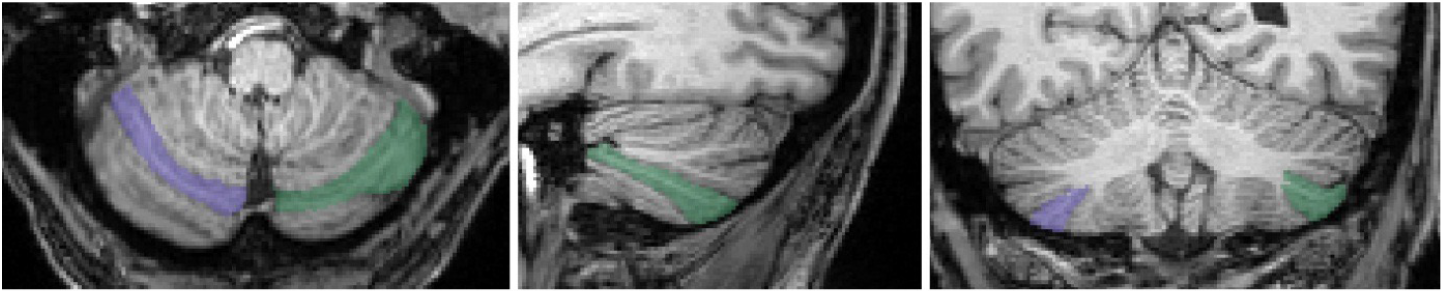
Lobule VIIB in axial, sagittal and coronal view. Green: left VIIB. Purple: right VIIB.

##### 2.4.7 Lobule VIIIA

Lobule VIIIA varies in the number of folia. In the midsagittal plane, the lobules VIIIA and VIIIB arise from the same branch of white matter, while further laterally the dichotomous branching is earlier recognizable. This is well recognizable mainly in the coronal view as well as in the sagittal view lateral to the midline (see Figure 13). In particular, the inferior parts of the segmentation need to be controlled in the axial view, since here the fissura intrabiventer is often well recognizable as a clear notch between the two lobules. See Figure 13.

**Figure 13:**
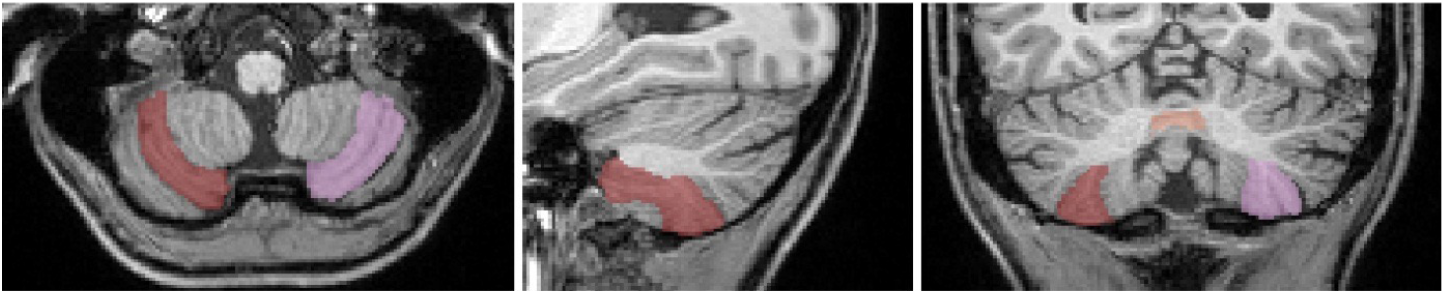
Lobule VIIIA in axial, sagittal and coronal view. Light pink: left VIIIA. Dark red: right VIIIA.

##### 2.4.8 Lobule VIIIB

The white matter branch from which lobule VIIIB originates often splits up in the periphery, see Figure 14, left picture. This subdivision can be easily identified in the axial view. In the sagittal view, lobule VIIIB curves from the outside around lobule IX, which lies more centrally. The fissura secunda, which separates lobules VIIIB and lobule IX, is identified and drawn in the coronal view as a relatively straight line. Partially one can identify a respective contrast between image intensities also in the sagittal view, allowing the separation of lobule VIIIB from lobule IX. In the anterior superior part, lobule VIIIB is also adjacent to lobule X; this boundary must be delineated in the sagittal plane. See Figure 14.

**Figure 14:**
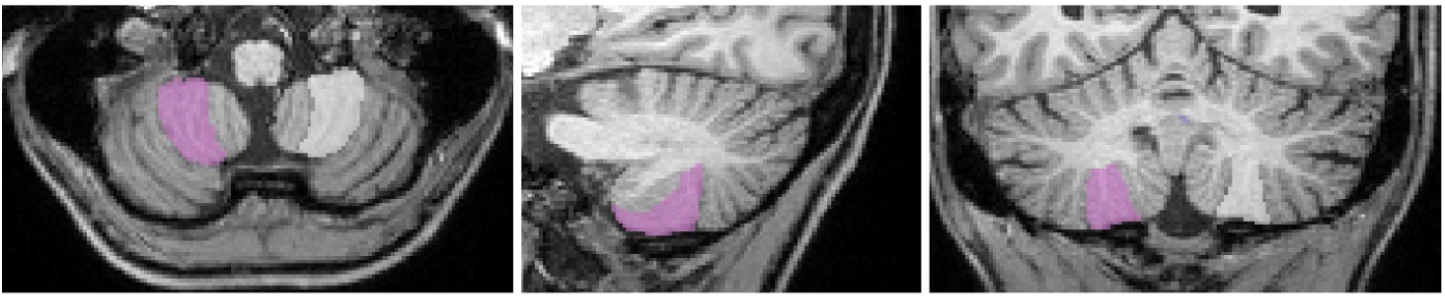
Lobule VIIIB in axial, sagittal and coronal view. White: left VIIIB. Pink: right VIIIB.

##### 2.4.9 Lobule IX

The left and right lobules IX are the most medial lobules and are adjacent to themselves as well as to the vermis in the midline. The borders of lobe IX are drawn in the coronal plane as almost straight lines. See Figure 15

**Figure 15:**
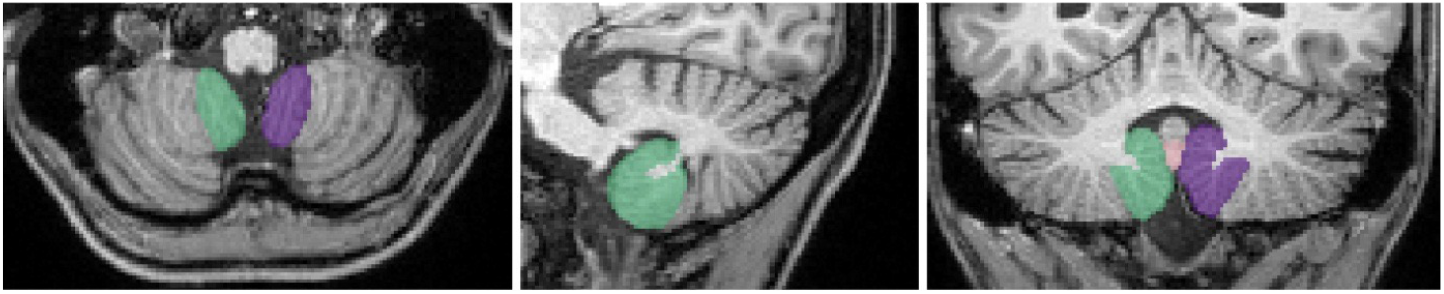
Lobule IX in axial, sagittal and coronal view. Purple: left IX. Green: right IX. Light pink: Vermis.

##### 2.4.10 Lobule X

Lobule X represents the flocculonodular lobe. It is delimited from lobule IX (Section 2.4.9) by the posterolateral fissure and can be located ventrally therefrom as a narrow structure. As mentioned above it also has contact with lobule VIIIB (Section 2.4.8. The fissure is easily recognizable in the lateral slices of the sagittal view and is drawn here. In principle, the outer boundaries of the cerebellum should already be correctly set in the first step (Section 2.1. However, we recommend doublechecking them when segmenting lobule X. In particular, accidental inclusion of cranial nerves should be checked in the axial view. See Figure 16, Figure 20.

**Figure 16:**
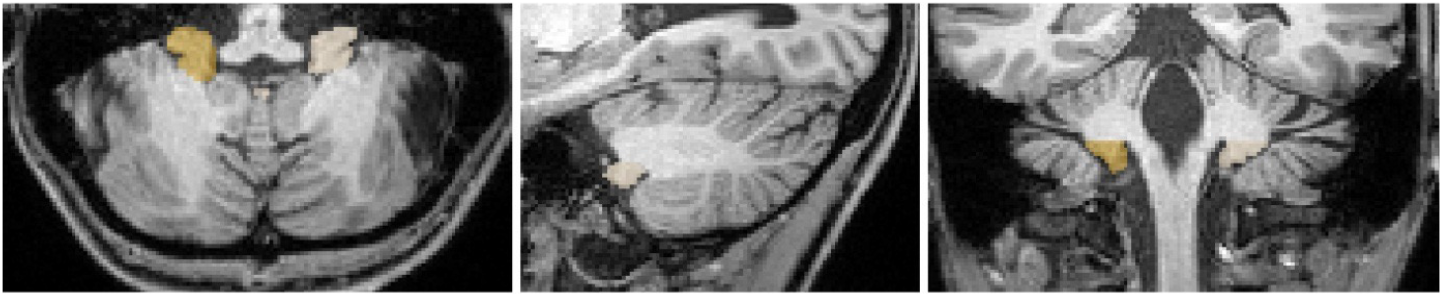
Lobule X in axial, sagittal and coronal view. Beige: left X. Ochre: right X.

#### 2.5 STEP 5: Sub-segmentation of the vermis

The already pre-defined entirety of vermis is now further subdivided corresponding to the hemispheric lobules VI, VII, VIII, IX and X, Figure 17. Clear idetification of fissures in the vermal or (para)midsagitall slices is often difficult. The subsegmentation is therefor oriented on both branches as well as fissures of the adjacent hemispheric lobules. The fissures bounding the lobules are tracked from lateral to medial and the delimitations within the vermis are drawn accordingly by extrapolating the medial completion of the lobule boundaries. Vermal sub-segmentation is oriented on the overall lobules: VI, VII, VIII, IX and X. A more fine-grained sub-segmentation would lead, at least in part, to a very small number of voxels. To identify Vermis VI, which impresses as an oblong structure between the two hemispheric lobules VI, the axial view is well suited.

**Figure 17:**
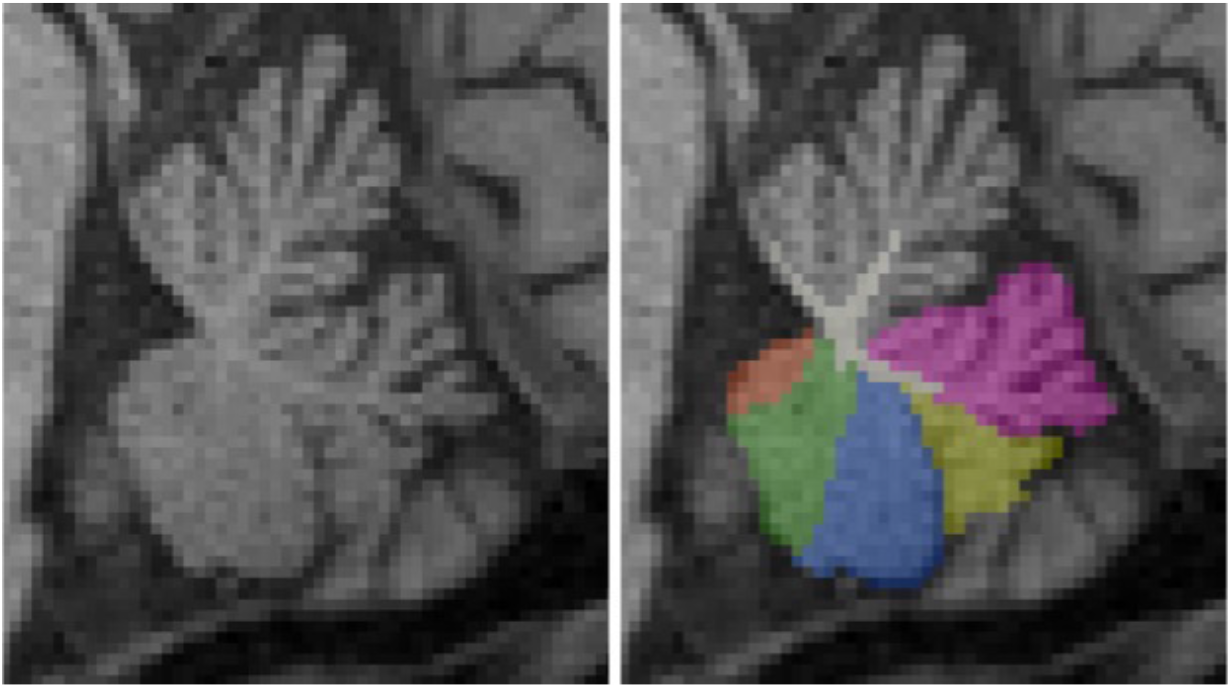
Sagittal view of the subdivided vermis corresponding to vermis VI (pink), vermis VII (yellow), vermis VIII (blue), vermis IX (green) and vermis X (orange).

In the more inferior parts, mainly the sagittal view is used to determine the border to Vermis VII. Vermis VII corresponds to the hemispheric lobule Crus I and II and VIIB. It is located inferior to Vermis VI. The more superior part, corresponding to the hemispheric lobules Crus I and Crus II is best seen in the axial and coronal view. Here, it is recognizable with a triangular shape. The adjacent fissures can be delimited in the midsagittal plane. The more inferior part, corresponding to the hemispheric lobule VIIB is identified in the axial view as a ventrally open, narrow arc. Vermis VIII occupies most of the vermis in the posterior-inferior cerebellum and is recognizable as a central circular structure in both the axial and coronal view. In the midsagittal plane the border to Vermis VII as well as Vermis IX is clearly visible and drawn in this view. Please note, that in the (mid)sagittal view an even more fine-grained sub-segmentation, corresponding to the hemispheric lobules VIIIA versus VI-IIB can be identified, but is not drawn. Vermis IX forms an arrowhead in the axial plane, which is framed by the Lobules IX of both cerebellar hemispheres. It is drawn in the midsagittal plane, where it has a relatively large extension. The boundary to Vermis X is best seen laterally of the midsagittal plane or in the axial view, which should therefore be used for doublechecking. Vermis X is the ventral tail end of the vermis and borders directly on the fourth ventricle. The posterolateral fissure between vermis IX and X is barely identifiable in the midline but is often more pronounced a few millimeters/slices further lateral in the sagittal view and can be medially completed by an imaginary extrapolating line from there. In general, for the vermal sub-segmentation all views need to be combined throughout the entire process of segmentation. See Figure 17.

#### 2.6 STEP 6: Segmentation of cerebellar white matter

##### GM/WM boundary towards the cerebellar cortex

The corpus medullare is defined as the white matter (WM) within the cerebellum and is connected to the brainstem via the three paired cerebellar peduncles. The deep cerebellar nuclei are located within the cerebellar WM, but are not definable on T1w MRI. Delineation of white matter branching to the finest ramifications that branch into the gray matter of the cerebellar cortex is limited due to the resolution and image contrast of the MRI. The pragmatic compromise that allows a consistent and reproducible approach is defined as follows. Segmentation of cerebellar WM branches starts in the midsagittal plane and is then continued in the sagittal view going laterally into the left and right hemispheres. Labeling starts in the corpus medullare towards the branches reaching into the cerebellar cortex, thereby correcting the provisory straight boundaries between the opposing limits of gray matter that were drawn in STEP1 (Section 2.2). Voxels that are categories by visual inspection of the image intensity as WM are labeled as white matter only as long as they share a common edge with the directly adjacent voxel that has already been assigned to white matter on the 2D view (corresponding to a common face in 3D). In exceptional cases, the isolated labeling of voxels that share only one corner in 2D and no edge in all views may be included if this ensures the overall continuity of a branch (Figure 18). Such cases should be carefully reviewed in all 3 views as well as the 3D model. In general we recommend to not only check but actively draw the white matter branches following the above mentioned rule in each view, in other words after going through the sagittal slices, branches of WM are checked and completed in the axial and coronal view. Finally, doublechecking the segmentation in all views as well as the 3D model.

**Figure 18:**
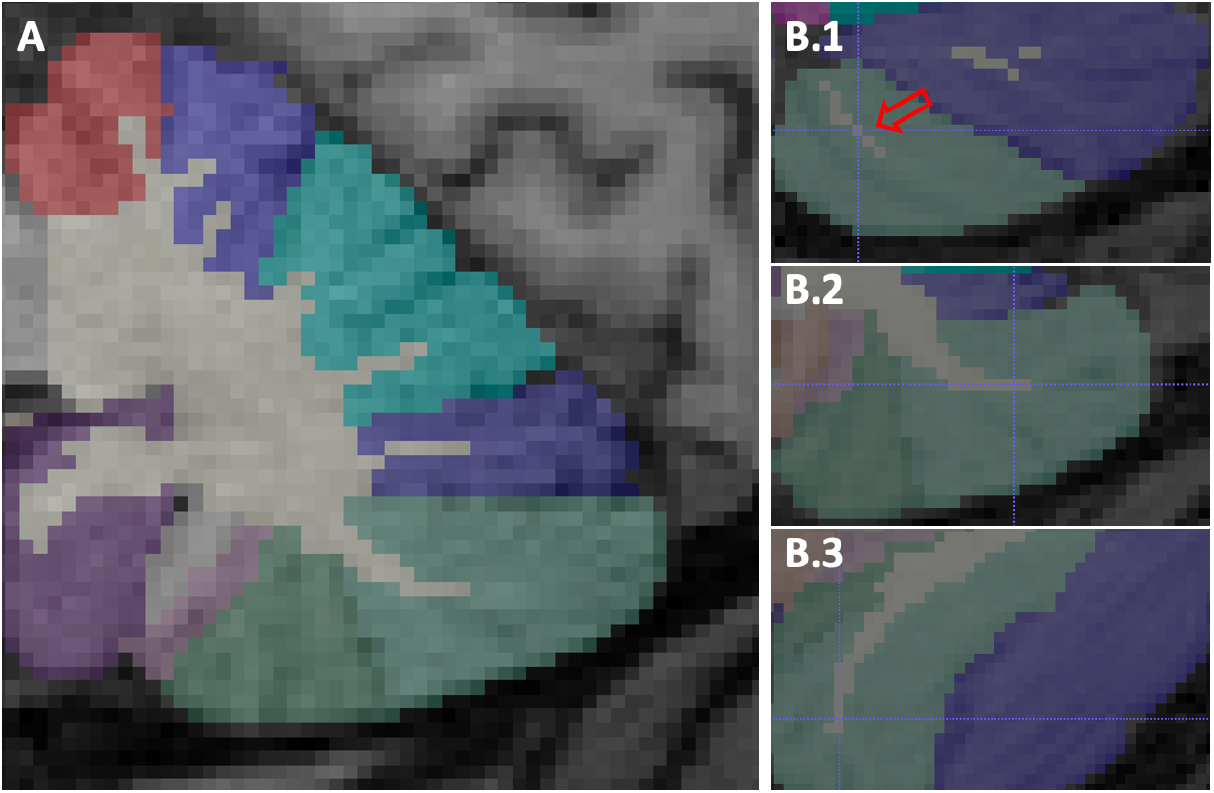
Segmentation of the cerbellar white matter with its branches reaching into the cerebellar cortex. Branches of white matter are drawn first in the sagittal view (A). Voxels visually identified as WM based on the image intensity are labeled as white matter as long as they share a common edge with a directly adjacent voxel that has already been assigned to white matter on a 2D plane (corresponding to a common face in 3D). In exceptional cases, the isolated labeling of voxels that share only one corner and no edge (like in the 2D example B.1) in all 2D views may be included if this ensures the overall continuity of a branch.

##### Boundary of cerebellar WM towards the brainstem

The cerebellum is connected with the brainstem, via the three paired cerebellar peduncles. Consequently, drawing a ‘boundary’ between cerebellar white matter towards the brainstem is somehow arbitrary and needs to be clearly defined to be consistent and reproducible. It is delineated in the sagittal view from one side to the other side and should be segmented as follows: First, in each sagittal slice, the connecting line between the superior and inferior point where crebellar cortex GM touches the WM is marked. Next, a direct, straight (linear) connecting line between those two points is drawn (Figure 19 A). The resulting boundary usually shows various “steps” that jump back and forth in the other two views. Thus, it needs to be corrected in the axial and coronal view to achieve a smooth outer surface. Please note, after correction in order to achieve a smoother surface, the boundary does not represent a straight/linear line in all sagittal views any longer. Finally, the smoothness of the outer surface should be checked in the 3D model (Figure 19).

**Figure 19:**
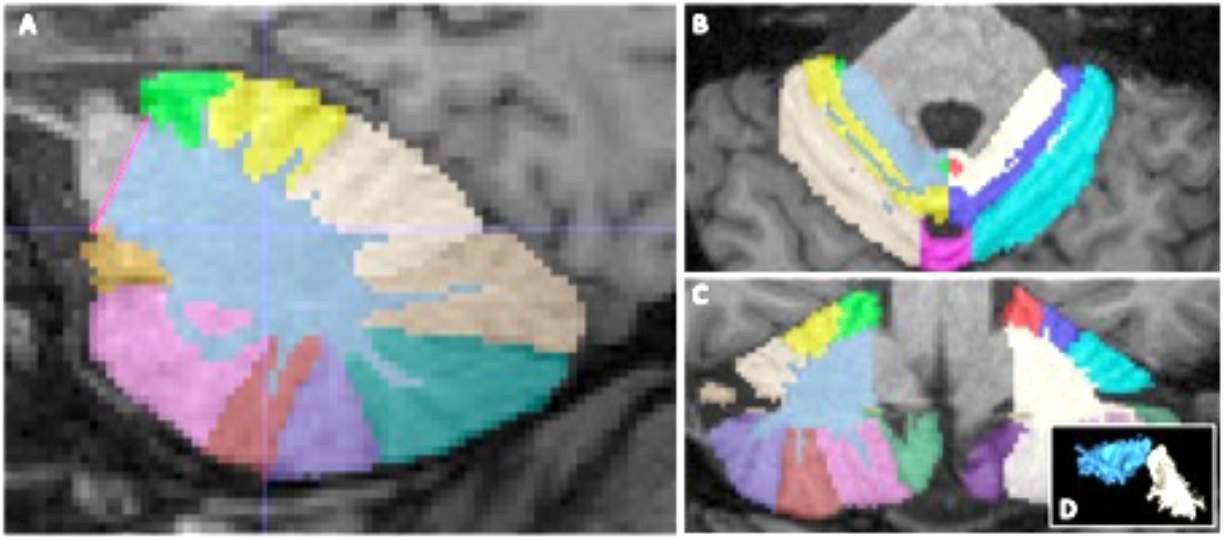
Segmentation example of the boundary of the cerebellar white matter towards the brainstem. First, in each sagittal slice, a connecting line between the superior and inferior point where cerebellar cortex GM touches the WM is drawn (A). Second, the boundary line is corrected in the axial (B) and coronal view (C) to achieve a smooth outer boundary. Third, the smoothness of the outer surface should be checked in the 3D model (D).

**Figure 20:**
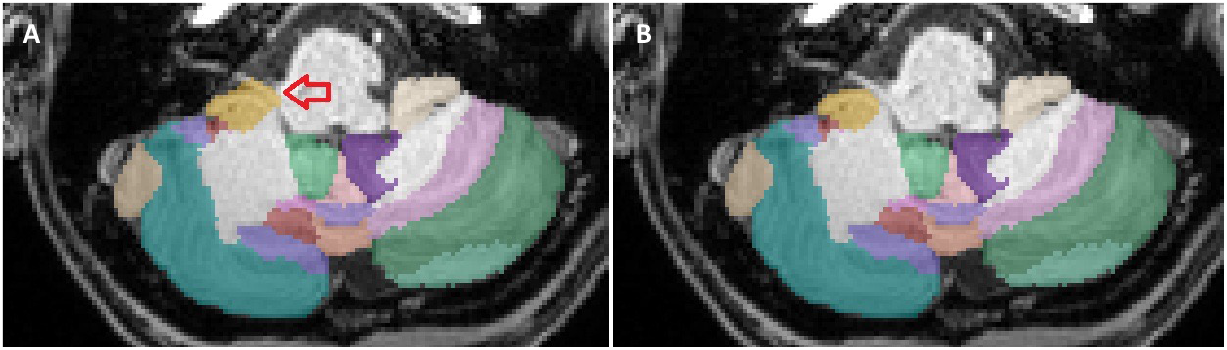
A cranial nerve that is located close to the cerebellum is marked on the left image by the red arrow (A). This misclassification was not identified in STEP 1. When labeling lobule X, this misclassification was found and subsequently corrected (B).

#### 2.7 Closing remarks

Ensure that each voxel is uniquely assigned to one cerebellar sub-structure of the *textbffinal* 27 cerebellar sub-structures:

*lobules I-IV, V, VI, Crus I, Crus II, VIIB, VIIIA, VIIIB, IX, X* and *cerebellar white matter, each left and right* as well as *vermis VI, vermis VII, vermis VIII, vermis IX, vermis X*.

Thus, resulting in a disjoint segmentation of the cerebellum. In other words, there should be no voxel labeled as for example “Cerebellar inferior posterior lobe” not assigned to either VIIIA, VIIIB, or IX.

Although copies of a segmentation can be useful as a backup, we generally recommend not to work with copies, but to consistently continue the hierarchical steps within one file.

Please note that the segmentation protocol does not take into account any phylogenetic or functional classifications of the cerebellum, such as pars median vs. pars intermedia and pars lateralis.

#### 2.8 Potential sources of misclassification recommendation how to solve them

Misclassification of cerebellar gray matter sub-structures can occur both at the outer border as well as between sub-segments within the cortex and do not delineate the exact outer demarcation of the cerebellum or the fissures recognizable in the MRI image. It can be either over-segmentations or under-segmentations omitting voxels that actually belong to the cerebellum. At the outer border of the cerebellum, the following structures may be misclassified as cerebellum: the tentorium, the most inferior parts of the cerebrum (Figure 4), the cerebral sinus/venes, and the cranial nerves (Example in Figure 20). Whereby the first mentioned are more likely to pose problems if automated pre-segmentations are used (Section 1.4.1. The latter however can also be problematic when drawing manually without pre-segmentations. Cranial nerves are in close proximity to lobule X and the distinction can be challenging especially in the sagittal view.

Thus, the axial plane must be used for crosschecking (as shown in Figure 20). As mentioned above, we recommend a hierarchical approach of segmentation; starting from large to more and more detailed sub-structures, by focusing at first only on the outer boundary of the cerebellum and use this overall segmentation as the ‘background label’ for the following sub-segmentation steps. By doing so, one avoids the erroneous inclusion of extracerebellar voxels in subsequent delineation steps. The correct assignment of sub-structures within the cerebellar cortex is much more challenging than the exact outer boundary. The main reasons are inclination and tilt of the folia and consequently the fissures in the three-dimensional space. Individuals may have divergent patterns of cerebellar lobule curvature, which can greatly affect the recognition of anatomical structures due to the unusual morphology, see Figure 22. These individual differences between brains make it difficult to find typical landmarks and thereby, can lead to erroneous assignments. Fissures are best recognized when they are exactly perpendicular to a 2D plane view and if their course is relatively straight and therefore easy to be tracked. However, this is in particular for the fissures between the lobules VIIB to VIIIB usually not the case (see also [10]). In order to achieve a satisfactory result, all vies must be considered and each structure iteratively corrected to achieve a consistently aligned, smooth and reasonable lobule structure. In any case of uncertainty, we strongly recommend to discuss the segmentations with colleagues and document the critical sub-strucutres.

**Figure 21:**
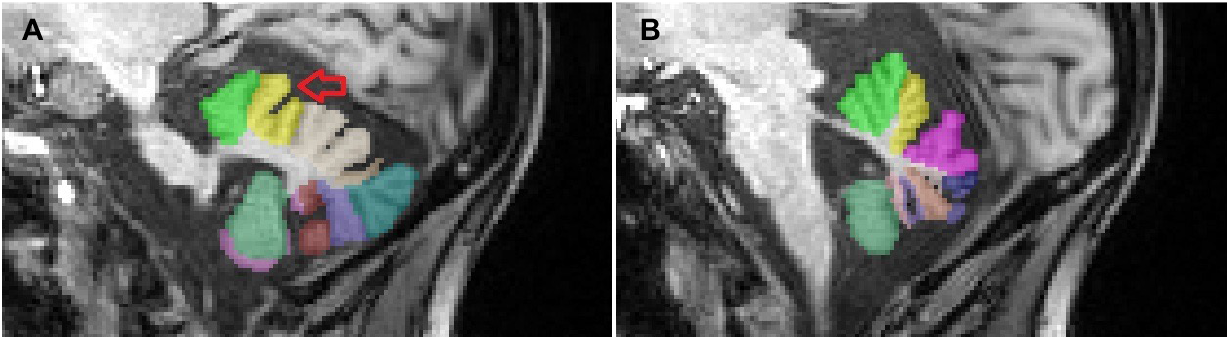
Cerebellum from the symptomatic patient, suffering from spinocerebellar ataxia type 3 (SCA 3) shown in the two sagittal planes. The image on the left shows a wide and relatively deep fissure (marked with a red arrow) within the right hemispheric lobule V (A). This fissure is no longer visible in the midline (B).

**Figure 22:**
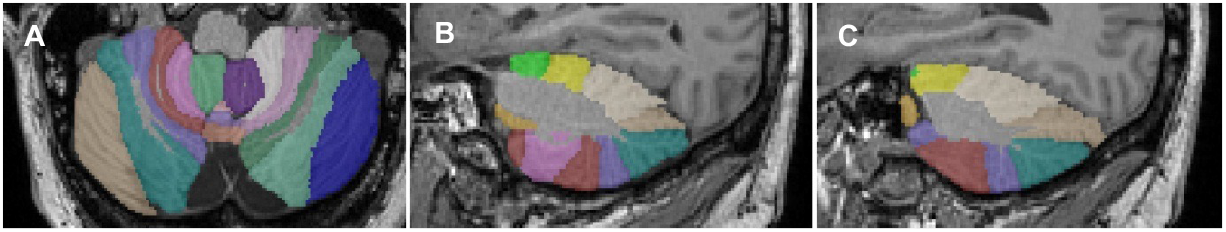
Cerebellum with pronounced curvature of the lobules VIIB to VIIIB. In the left picture the cerebellum is shown in one axial slice (A), in the middle and right picture in two different sagittal slices (B, C). The curvature changes the morphology so that the same lobule (marked in purple) appears ‘twice’ in one plane (C).

Another source of error, are MR image intensity changes that are misinterpreted as fissures. We recommend to document the image setting in advance and doublecheck all structures in all 3 views. Finally, the high inter-individual variability of the cerebellum is a common source of error. Deviations from the expected ‘anatomical standard’ are common, such as e.g. the number of folia of one lobule. In such cases, the correct assignment of each folium to one or the adjacent lobule is crucial. This can result in two different ‘interpretations’ of one folium that is assigned to the one or the other, neighboring lobule. A challenge that one is frequently confronted with, when searching for the the superior posterior fissure, in particular if an additional folium of Crus I or lobule VI is present, that does not reach the periphery. In general, we recommend to go back to the white matter branches and search for/track the connections to the respective WM of the particular lobules. However, these branches are sometimes very narrow and might only be visible in one view. Therefor cross-checking all views, again, is essential. If no fissures are visible at all on the image, the lobules should be derived entirely from the branches of the white substance. Also be aware, that some intra-lobule fissures Figure 21 might appear even deeper and larger than the fissures that actually separate two lobules (see also [10]). For example, segmentation of substructures in an atrophied cerebellum, such as in an ataxia patient, can be challenging because although the fissures are wider and deeper due to atrophy, many of the intra-lobular fissures are prominent that are not normally seen in healthy brains. Subsequently this can lead to uncertainty in the correct labeling. Again, in case of uncertainties regarding the correct assignment of the lobules, going back to the corpus medullare and following its branching out is helpful.

In order to avoid misclassifications in the assignment of the labels, we recommend to cross-check every drawn structure and its delimitations directly afterwards and the final sub-segmentation in all three views and the 3D viewer in addition. One should take notes of all uncertainties in the documentation file and have a second look after some time, ideally together with a (neuro)radiologically experienced colleague.

## Data Availability

All data produced in the present study are available upon reasonable request to the authors.

## 3 Acknowledgements

We would like to thank Beate Brol, Tim Elter, Isabelle Finkel, and Sophia Wismeth for their contribution to the manual segmentation.

This work was supported by the National Ataxia Foundation’s SCA Young Investigator Award.

## 4 Appendix

### Comparison to other segmentation protocols

Park and colleagues [11] and Bogovic and colleagues ([10], with more detailed information that can be found here: http://iacl.ece.jhu.edu/index.php?title=Protocol_for_Cerebellar_Labeling) have manually segmented the cerebellum. In the following sections we will briefly comment on both approaches.

#### Segmentation of the boundary between cerebellar white matter and the brainstem

While Bogovic et al. have chosen to associate the inferior interface of gray and white matter and cerebrospinal fluid with the superior interface like we do, Park et al. defined a virtual line, which (a) is orthogonal to the AC-PC connection (anterior to posterior commissure) and (b) touches the posterior border of the inferior colliculi. It is defined in the midsagittal plane and then continued to all other sagittal slices.

#### Segmentation of the boundary between cerebellar white matter and cerebellar cortical gray matter

Bogovic et al. as well as Park et al. do not segment any branching ramification of white matter into the cerebellar cortex like we do but rather just stops at the bottom of any folia.

#### Sub-segmentation of the anterior lobe

Park et al. subsegment lobules I-II, III, IV and V in the anterior lobe, while Bogovic et al. segment I-III, IV and V. Lobule III has a semilunar form and in most cases the first distinct folium not attached to the superior medullary velum. However it is often at least partially obscured by lobule IV. Like in the spatially unbiased atlas template of the cerebellum and brainstem, SUIT, [4] we also decided not to subdivide lobules I-IV for now, due to the partially vague fissures together with the close functional context (Section 1.3).

#### Sub-segmentation of vermis

Park et al. delineated the individual lobules, but combined the hemispheres and the vermis to simplify the anatomy and avoid a possibly arbitrary limitation. In contrast, Bogovic et al. segmented the midline vermis in their publication corresponding to the hemispheric lobules VIIIA, VIIIB, IX and X. The segmentation of the vermis corresponding to the lobules VI-VII is commented with “under construction” on their webpage. The automated method for cerebellar subsegmentation, ACAPULCO [14], that refers to the work of Bogovic et al. does cover the vermal sub-segments: VI, VII, VIII, IX and X. Like ACAPULCO our protocol includes a sub-segmentation of the vermis into vermis VI, VII, VIII, IX and X corresponding to the respective hemispheric lobules.

#### Segmentation of deep cerebellar nuclei

Park et al. have segmented the deep cerebellar nuclei on the T2w MRI that were acuqired in addition to T1w scans. Like Bogovic et al. our segmentations are only based on T1w MRI, where the deep cerebellar nuclei cannot be identified.

#### Hierarchization of segmentation

Park et al. segment the lobules sequentially, beginning with lobule I to lobule X. In contrast, Bogovic et al. focused initially on larger structures going to more detailed sub-structures, like we do. In our opinion such a hierarchical process has the advantage, that the most prominent fissures that subdivide the lobes can be easier located and thereby already define unquestionable landmarks and provide a clearer structure.

### Overview of segmented cerebellar sub-structures in different protocols as well as for different automated methods

The following table gives an overview of segmented sub-structures of the cerebellum in different segmentation protocols (Park et al. [11], Bogociv et al. [10] and the protocol presented here (Faber et al.) as well as for different methods for automated cerebellar subsegmentation(SUIT [4], CERES [15, 16], ACAPULCO [14] and CerebNet).

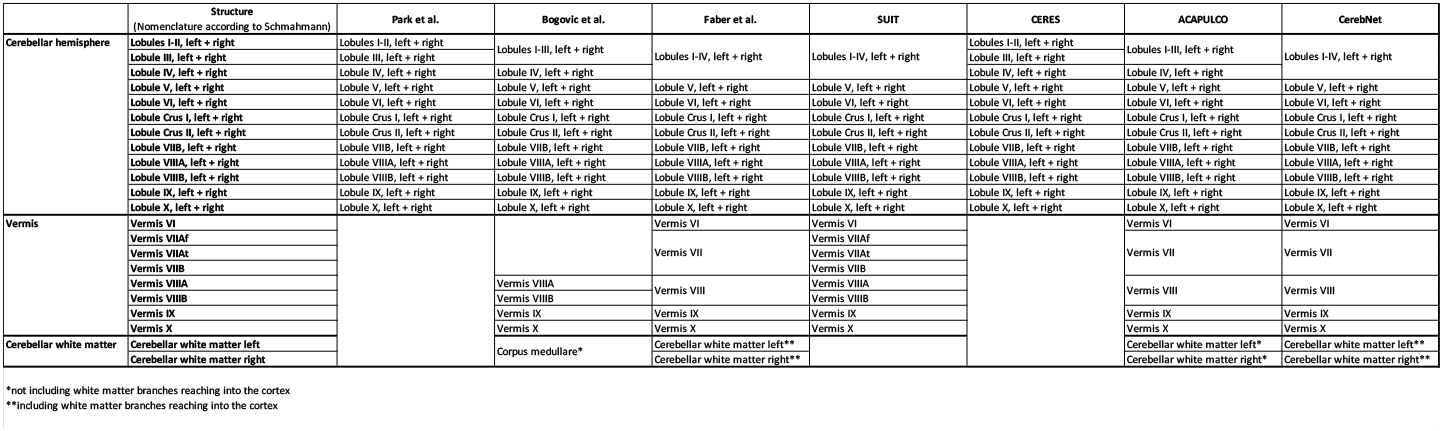

